# Sample Adequacy Controls for Infectious Disease Diagnosis by Oral Swabbing

**DOI:** 10.1101/2020.07.17.20156489

**Authors:** Meagan Deviaene, Kris M. Weigel, Rachel C. Wood, Angelique K. K. Luabeya, Lisa Jones-Engel, Mark Hatherill, Gerard A. Cangelosi

## Abstract

Oral swabs are emerging as a non-invasive sample type for diagnosing infectious diseases including Ebola, tuberculosis (TB), and COVID-19. To assure proper sample collection, sample adequacy controls (SACs) are needed that detect substances indicative of samples collected within the oral cavity. This study evaluated two candidate SACs for this purpose. One detected representative oral microbiota (*Streptococcus* species DNA) and the other, human cells (human mitochondrial DNA, mtDNA). Quantitative PCR (qPCR) assays for the two target cell types were applied to buccal swabs (representing samples collected within the oral cavity) and hand swabs (representing improperly collected samples) obtained from 51 healthy U.S. volunteers. Quantification cycle (Cq) cutoffs that maximized Youden’s index were established for each assay. The streptococcal target at a Cq cutoff of ≤34.9 had 99.0% sensitivity and specificity for oral swab samples, whereas human mtDNA perfectly distinguished between hand and mouth swabs with a Cq cutoff of 31.3. The human mtDNA test was then applied to buccal, tongue, and gum swabs that had previously been collected from TB patients and controls in South Africa, along with “air swabs” collected as negative controls (total N = 292 swabs from 71 subjects). Of these swabs, 287/292 (98%) exhibited the expected Cq values. In a paired analysis the three oral sites yielded indistinguishable amounts of human mtDNA, however PurFlock™ swabs collected slightly more human mtDNA than did OmniSwabs™ (p = 0.012). The results indicate that quantification of human mtDNA cannot distinguish swabs collected from different sites within the mouth. However, it can reliably distinguish oral swabs from swabs that were not used orally., which makes it a useful SAC for oral swab-based diagnosis.

## INTRODUCTION

Oral swabbing is a non-invasive, inexpensive, and easy-to-use sampling strategy for infection diagnosis. An oral swab sample is collected by brushing a swab on a surface within the mouth, such as the tongue dorsum or buccal mucosa (inner cheek). Oral swabs have been employed in experimental or clinical contexts to detect Ebola virus, human papillomavirus, *Mycobacterium tuberculosis, Leishmania*, the novel coronavirus SARS-CoV-2, and other infectious diseases in both humans and animals. (1–11) Compared to more traditional specimens such as sputum, urine, blood, and nasopharyngeal swabs, oral swabbing is faster, less invasive, and requires no accommodations for privacy or aerosol control. In the case of TB, recent studies showed that qPCR analysis of oral swabs can detect *Mycobacterium tuberculosis* DNA in up to 93% of sputum qPCR-positive adult pulmonary TB patients. (1,2,8) One of these studies showed that tongue swabs yield significantly better *M. tuberculosis* signal than cheek or gum swabs. (1) In another recent study, self-collected tongue swabs detected 47/50 patients with confirmed COVID-19, for a sensitivity of 90% relative to the more invasive nasopharyngeal swabbing method.(3) Such results illustrate the expanding potential for simple swabbing methods in the molecular diagnosis of infectious diseases, and the need for additional research in this area.

One hindrance to the evaluation and implementation of oral swab methods is the potential for false negative results due to inadequate sampling (insufficient contact with oral mucosa). This problem is especially acute in the context of self-sampling as envisioned for scaled-up COVID-19 screening.(3,12) In contrast to samples such as sputum and blood, which can be inspected visually to assure collection, samples collected within the mouth are most often visually indistinguishable from those which have never been inside a mouth. This challenge can be met by using a sample adequacy control (SAC), defined as a molecular target which differs from the diagnostic target and should consistently be present in definable amounts in properly collected specimens.

Though standards for evaluating diagnostic tests frequently include quality assurance (QA) strategies (13–15) components and levels of QA are differentially adopted. Often, reference materials are added to samples as positive controls, for example purified DNA (16). These are good practices to validate laboratory processes but they do not address the upstream issue of sampling adequacy. Even among approved molecular diagnostic assays, use of sample adequacy controls is not uniform. (17) In some cases, SACs are successfully utilized without preliminary validation. For example, in recent reports on the successful use of oral swabbing and virus nucleic acid testing for Ebola virus, endogenous controls targeting human nucleic acids were used as SACs. However, no determination of the amount of such nucleic acids that constitutes a swab collected within the oral cavity were presented. (4,5)

The present study evaluated two qPCR assays for their potential use in verifying whether a swab has sampled material within the mouth. One candidate SAC exploited the expanding understanding of the human oral microbiome in order to design a qPCR test specific for some of these microbes. The oral cavity has a diverse microbiota, with over 700 species identified, (18) yet among human microbiomes it has one of the lowest levels of beta diversity. (19) *Streptococcus* is a core genus of human oral microbiomes (20) and one of our candidate SAC tests targeted species of this genus.

The second candidate SAC test targeted a ubiquitous human mitochondrial gene, under the assumption that swabs with sufficient contact with the oral epithelium will consistently contain more human cells than swabs which had been handled but not had contact with the oral cavity. Polymorphisms within mtDNA genes are rare and mitigated by the heteroplasmy of mtDNA. (21,22) Housekeeping genes, including mtDNA genes, are often used as SACs in infectious disease diagnostic tests. Most often this has been to ensure that inhibition is not occurring in the qPCR amplification, rather than for the purpose of ascertaining sampling adequacy. (16,23,24)

To validate the candidate SACs, a study was undertaken that was paired in two ways. First, hand and mouth swabs were tested from each individual. Second, two qPCR assays (for *Streptococcus* species and for human mtDNA) were performed on each sample. Hand swabs were used as simulated mishandled swabs that incidentally contacted human skin outside of the mouth. Such swabs were expected to contain both human and streptococcal DNA, but in smaller amounts than mouth swabs. We tested the hypothesis that mouth swab Cq values are sufficiently different from hand swab Cq values to enable the establishment of a threshold value separating the two groups. After optimization, a method utilizing human mtDNA was then applied to oral swabs collected by diverse methods from a cohort in Worcester, South Africa, as part of a recently completed analysis of oral swab testing for testing for TB. (1)

## MATERIALS AND METHODS

### Sample populations

Subjects in Seattle, WA, USA were recruited via flyers and emails that were posted and sent throughout the University of Washington campus. Samples were collected from subjects in their workplaces. Samples were stored at −80 °C within 8 hours of collection, and subsequently tested by qPCR as described below.

Samples archived as part of a previous study on the use of oral swabs in the diagnosis of adult pulmonary TB patients in South Africa (1) were also used for the current study. South African participants had been recruited in clinics in Worcester, Western Cape (population 350,000). As described previously, these samples were collected from subjects in clinics or homes, stored at −80 °C within 8 hours of collection, and shipped frozen to Seattle where they were assayed by qPCR for *M. tuberculosis* DNA. (1) For the current study these archived samples were screened for human mtDNA.

Protocols for collecting samples from U.S. subjects were approved by the University of Washington’s Human Subjects Division. Archived South African samples collected for the previous study focusing on TB diagnosis (1) were used in the current study under “future use” provisions in consent forms, as approved by the University of Washington and the University of Cape Town.

### Swab Collection

In Seattle, samples were collected by brushing Whatman OmniSwabs™ along the inside of each subject’s cheek (oral swabs) or the surfaces of the palm and fingers (hand swabs) 7-8 times, or about 5 seconds.

In South Africa, OmniSwabs™ and Puritan PurFlock™ swabs were previously collected by brushing the inner surface of the tongue dorsum, gums, or cheeks for 10 seconds. (1) “Air controls” were collected at both study sites by exposing a swab to air for 10 seconds (PurFlock™ in South Africa, OmniSwab™ in Seattle). Immediately following sampling, all swab heads were deposited into 500 mL of a lysis/preservation buffer (pH 8.0) consisting of 50 mM EDTA and sucrose, 100 mM sodium chloride, 65 mM tris, and 0.3% SDS. Samples were stored frozen at −80 °C until use.

For the present study a total of 292 South African swabs were tested. They were selected based on the availability of partially processed samples, at a time point in the previous study (1) when TB status was blinded to us. The 292 swabs came from 71 South African subjects, each of whom provided up to 5 swabs (4 oral and 1 air). Some subjects had fewer than 5 swabs tested for this study. Only subjects for whom all 4 oral swabs were tested (N = 32) were included in comparisons between swabbing sites and swab brands. After unblinding, this cohort was found to include 7 TB-positive subjects and 25 TB-negative subjects.

### DNA extraction

Samples collected from U.S. volunteers were extracted using the QIAGEN 96 DNeasy Blood and Tissue Kit protocol. To begin extraction samples were boiled in lysis/preservation buffer at 100 °C for 10 min, followed by addition of proteinase K and QIAGEN lysis buffer AL and a 10-min incubation at 56 °C. Ethanol was then added to the samples and they were transferred to a plate column well for binding, two washes and elution. Nucleic acid was extracted from swab samples collected in South Africa using a modified spin column protocol of the QIAGEN QIAamp DNA mini kit (catalog #51306), described previously (1).

### Quantitative PCR

Each U.S. sample was tested twice, once for streptococcal DNA and once for human mtDNA (Table 1). The *Streptococcus* species qPCR was adapted using primers from Picard et al. (25) with the addition of a hydrolysis probe specific for four species of *Streptococcus*, namely *S. gordonii, S. mitis, S. pneumoniae*, and *S. sanguinis*. These species were selected because of their established presence in the oral microbiome, combined with a limited known presence on the skin or in the environment, as reported by the Human Microbiome Project (hmpdacc.org/HMRGD). The buffer and primer concentrations were identical to the published protocol, (25) with the modifications of probe concentration at 0.5 µM in the final reaction mixture and 2 µL of template instead of 1 µL. The human mtDNA qPCR was adapted from Caldwell et al. (26) qPCR primer and probe sequences as well as thermocycling conditions are summarized in Table 1. For the samples from South Africa, 5 µL of archived extracted DNA from the previous study (1) was tested only by the assay targeting human mtDNA.

**TABLE 1.**
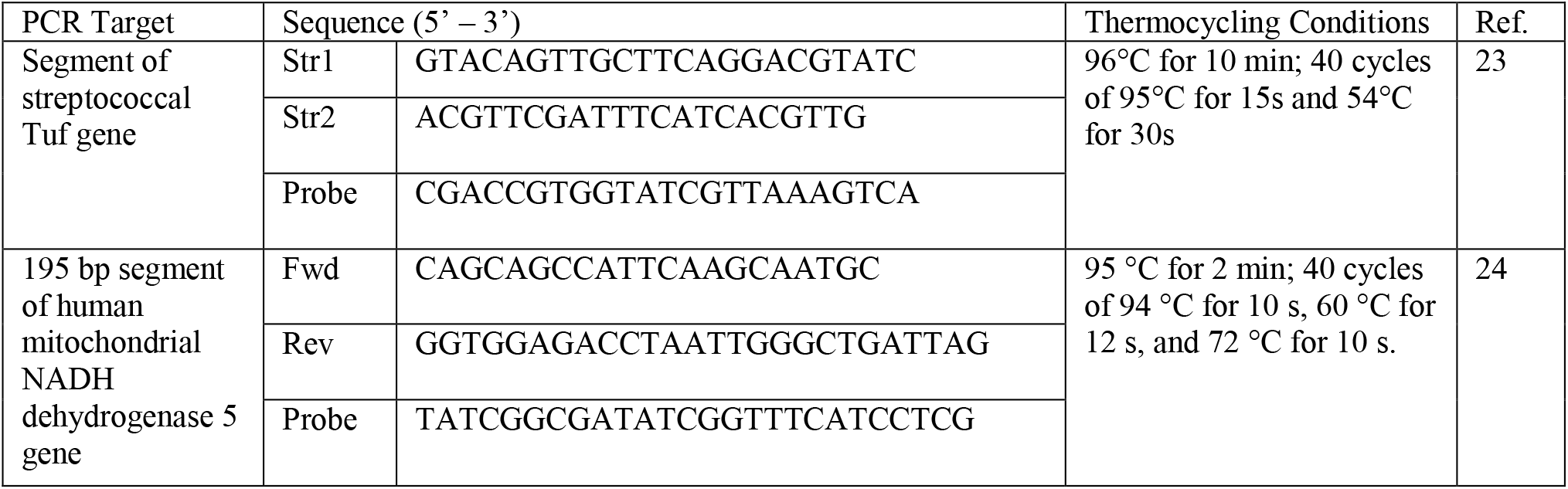
PCR technical information.

### Statistical Analyses

Calculations were carried out using R, version 3.1.3. The package OptimalCutpoints was used to calculate Cq cutoffs with Youden’s index. T-tests for both Seattle and South African studies were performed on paired samples because they evaluated within-subject differences.

## RESULTS

### Comparison of two SAC targets

We first compared streptococcal DNA and human mtDNA as markers for oral swab SAC. While both markers were assumed to be ubiquitous on human skin and in the environment, we hypothesized that swabs with sufficient contact with the oral cavity would consistently have more of these materials than swabs that had not been inside human mouths. To test this, 51 healthy U.S. subjects were enrolled and 5 OmniSwabs™ were collected per participant: One from each hand, one from each inner cheek, and one “air control” swab, which constitutes a swab exposed to air for 10 seconds to serve as a negative control. Cq values (inversely proportional to target quantity) from streptococcal and human mtDNA qPCR tests conducted on all 255 samples (n = 510) were compared.

Results from qPCR analyses of mouth and hand swabs are summarized in Figure 1. A Cq cutoff value that maximized Youden’s index ([sensitivity + specificity] − 1) was established for each assay. The qPCR assay for streptococcal DNA had an optimal Cq cutoff at 34.9. Cq values below this cutoff corresponded with oral swabbing with a sensitivity of 99.0% and a specificity of 99.0%. Thus, the streptococcal assay was imperfect as a SAC. In contrast, human mtDNA as a classifier was able to perfectly distinguish between hand and mouth samples with a threshold Cq value of 31.3 (Figure 1). Air swabs yielded no detectable qPCR signal for either human mtDNA or streptococcal DNA in 94% of cases (48/51). When qPCR signals were observed in air swabs, their Cq values were always above the optimal cutoff value.

**Figure 1.**
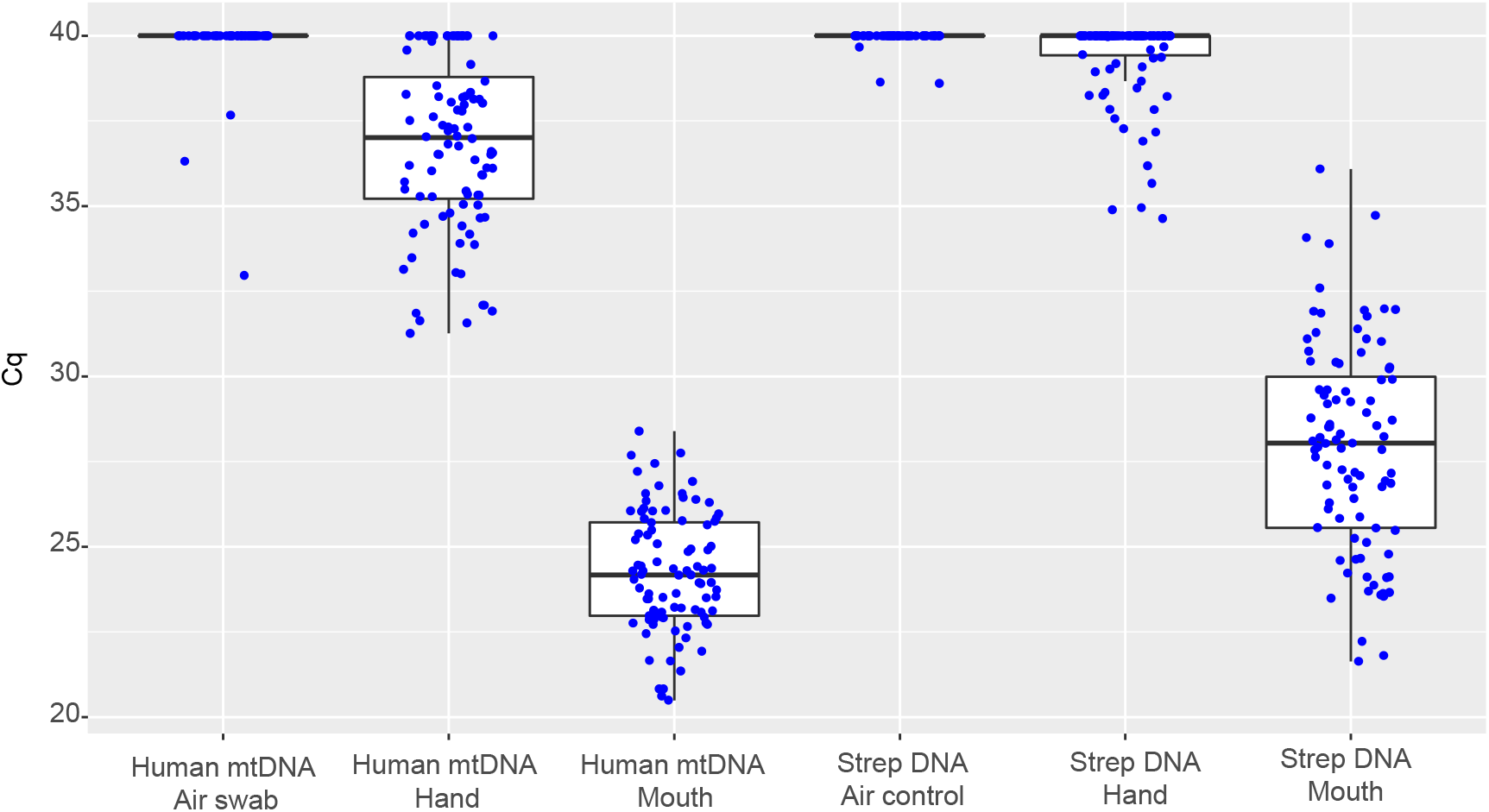
Oral swabs, hand swabs, and air swabs collected in the US. Data points are Cq values (inversely proportional to target DNA quantity) from qPCR analyses of cheek, hand, and air swab samples from 51 healthy U.S. subjects. Each sample was tested for streptococcal (Strep) DNA and human mtDNA. Samples with no detectable qPCR signal were assigned Cq values of 40.

### Comparison of oral sites and swab brands

Next, the human mtDNA sample adequacy control was applied to samples previously collected in a blinded study from South African TB patients and controls. (1) In Phase 1 of the previous study, South Africans with possible TB sampled by oral swabbing. Swabs were collected from each subject on three different days. For the present study, a total of 292 swabs from 71 South African subjects were tested for human mtDNA. The oral site and swab brand comparisons focused on oral swabs collected from 32 participants during their second swabbing session (Day 2). The swabs were collected from three different oral sites using two different swab brands, specifically: Tongue dorsum (OmniSwabs™), gum (OmniSwabs™), right buccal surface (OmniSwabs™), and left buccal surface (PurFlock™). (1)

Cq values from buccal OmniSwab™ samples collected from U.S. and South African donors (Figures 1 and 2, respectively) were compared in a T-test for differences in human mtDNA. Buccal OmniSwab™ Cq values from the South African cohort were significantly lower (H_A_ = two-sided) than those from the initial evaluation done on the U.S. cohort (p=0.0016). This difference may reflect disparities in sampling or DNA extraction methods. Swabbing duration for South African participants was 10 seconds, longer than for the participants in the Seattle phase of this study (5 seconds). Different QIAGEN extraction kits were utilized for samples deriving from the different sites.

**Figure 2.**
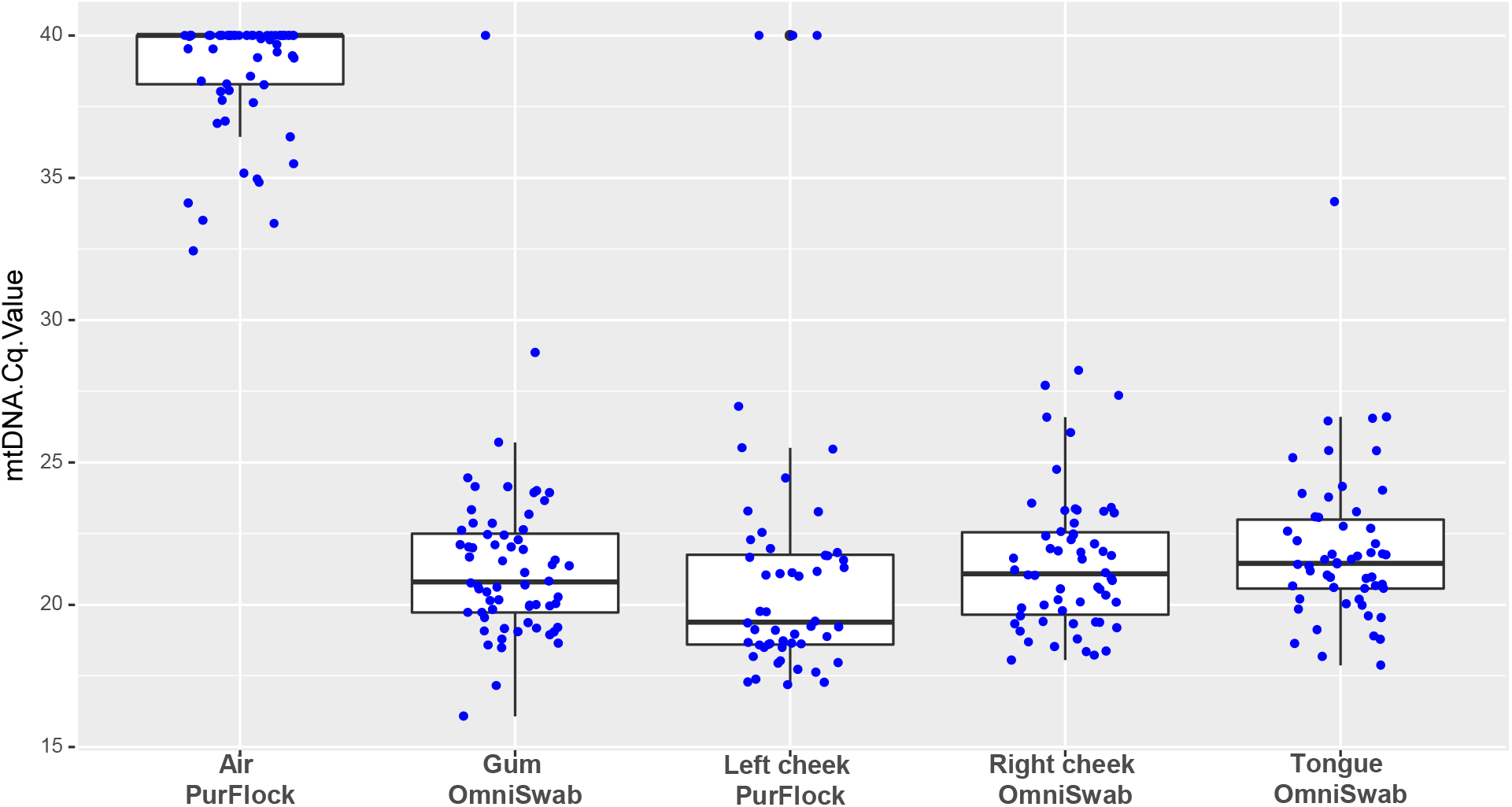
Oral swabs and air swabs collected in South Africa. Data points are Cq values (inversely proportional to target DNA quantity) from qPCR analyses of cheek, gum, and tongue samples and ambient air swabs collected in South African sites from 71 South African subjects. All sites were sampled with OmniSwabs except left cheek (PurFlock). Each sample was tested for human mtDNA. A subset amples for which all 5 sample types were collected were compared by paired T-tests for differences in mtDNA yield.. Samples with no detectable qPCR signal were assigned Cq values of 40.

Comparing mouth regions, in a paired analysis Cq values for gum or tongue OmniSwabs™ were not significantly different from buccal OmniSwabs™ (Figure 2). Comparing swab types, Purflock™ buccal swabs yielded lower Cq values (indicating more human mtDNA) than OmniSwab™ buccal swabs (p=0.019) (Figure 2). Thus, it appears that PurFlock™ swabs collect slightly more human biological material than OmniSwabs™ in the same mouth region.

### Human mtDNA in ambient air

The South African sample did not include hand swabs, however it did include PurFlock™ “air swabs” that were collected in South African homes and clinics at the same time that oral swabs were collected. The human mtDNA test was applied to a sample of 64 of these swabs, 20 of which were collected at the same time and location as the oral swabs in Figure 2. Human mtDNA was detected in many of these swabs. However, Cq values always fell above the threshold value of 31.3, indicative of orally collected swabs (Figure 2).

## DISCUSSION

Oral swabs are emerging as non-invasive samples for diagnosing infectious diseases. In the most sensitive protocol identified to date for oral swab diagnosis of TB, (1) the dorsum of the tongue is gently scraped and the swab head with collected material, consisting of bacterial biofilm and host cells, is deposited into a sample buffer; the sample is a semi-clear, non-viscous suspension suitable for testing by qPCR targeting *M. tuberculosis* DNA. Tongue swabbing is fast, painless, and does not require accommodations for privacy or aerosol control. Sputum-scarce patients such as children and HIV-positive adults are easily swabbed. In recent studies the method approached the sensitivity and specificity of sputum testing in adults. (1) In a separate study, the method approached the sensitivity and specificity of gold-standard nasopharyngeal swabbing for the detection of COVID-19. (3)

Despite these advantages, additional work is needed to bring the method into parity with traditional methods. For example, in one study buccal swabs collected from TB patients exhibited only 45% sensitivity relative to sputum testing. (8) In that study swabs were tested using the Cepheid GeneXpert Ultra using a protocol designed for sputum (not swab) sample processing and testing. Moreover, it used buccal swabs, (2) not the more optimal tongue swabs. (1) Such results support the feasibility of oral swabbing for TB, but highlight the need for more research to yield fully optimized methods. As such work proceeds, it is essential to control for possible false negative results that arise from inadequate sampling. In contrast to visually distinctive specimens such as sputum and blood, a swab collected in the oral cavity can resemble a swab that has never been inside a mouth. Therefore, the current study identified and evaluated SACs for use in evaluating and implementing oral swab-based diagnostic methods.

While tests targeting human mtDNA and streptococcal DNA both appeared able to distinguish mouth swabs from hand swabs, the human mtDNA test was more sensitive and specific at a calculated Cq cutoff. This was largely due to the more variable and higher overall Cq values observed when mouth swabs were tested by the streptococcal assay. This observation is consistent with the beta diversity that has been reported for human oral microbiomes. (21–23)

It is noteworthy that a significant number of air swabs had positive signals for human DNA, albeit well outside the threshold Cq value. The number was larger in the South African sample than in the U.S. sample, possibly because samples were collected in more diverse environments in South Africa, including clinics and patient homes. Human DNA is ubiquitous in built environments. (27) This underscores the need to use quantitative methods combined with a well-validated threshold value when human DNA is used as a SAC.

All U.S. oral swab samples fell below the human mtDNA Cq threshold for orally collected swabs. In the South African sample, 5/292 oral swabs (1.71%) fell above the Cq threshold established in this study. This included 3 buccal swabs, one gum swab, and one tongue swab. These samples were excluded from the previous study. (1)

The human mtDNA qPCR appears to be robust and useful as a SAC for determining whether a swab has been collected in several mouth regions using 2 swab brands. Nonetheless, some quantitative variations were observed. The South African samples had lower Cq values (more human mtDNA) than the U.S. samples. This might reflect differences in sampling methods, prevalence of TB, and/or comorbidities, diet, socioeconomic factors, and/or microbiomes.

This study had several limitations. The sample size was limited and restricted to two geographical locations in the U.S. and South Africa. The South African sample was a convenience sample consisting of people visiting a clinic for TB-like symptoms. Although it included air swabs, it did not include hand swabs. The current analysis was non-blinded. Neither human mtDNA nor Streptococcal DNA were found to be useful for distinguishing swabs collected from different sites within the mouth.

Despite these limitations, the results indicate that PCR for human mtDNA can reliably distinguish oral swabs from swabs that were not used orally. This feature makes it a useful SAC for oral swab-based diagnosis. The human mtDNA SAC may prove useful in the evaluation and implementation of oral swabs as non-invasive tools for infectious disease diagnosis.

## Data Availability

Data are provided in the article. Additional data available upon request

## ACKNOWLEDGEMENTS

The authors wish to thank Felicia Nguyen and Alaina Olson, who aided in sample analysis, and volunteers who provided samples for this project. We are also grateful to the staff at SATVI in South Africa for their role in recruiting and sampling participants at that site. This work was supported by a Grand Challenges Explorations grant (OPP1191195) from the Bill & Melinda Gates Foundation.

